# A pandemic at the Tunisian scale. Mathematical modelling of reported and unreported COVID-19 infected cases

**DOI:** 10.1101/2020.05.21.20108621

**Authors:** Ines Abdeljaoued-Tej

## Abstract

Starting from the city of Wuhan in China in late December 2019, the pandemic quickly spread to the rest of the world along the main intercontinental air routes. At the time of writing this article, there are officially about five million infections and more than 300 000 deaths. Statistics vary widely from country to country, revealing significant differences in anticipation and management of the crisis. We propose to examine the COVID-19 epidemic in Tunisia through mathematical models, which aim to determine the actual number of infected cases and to predict the course of the epidemic. As of May 11, 2020, there are officially 1032 COVID-19 infected cases in Tunisia. 45 people have died. Using a mathematical model based on the number of reported infected cases, the number of deaths, and the effect of the 18-day delay between infection and death, this study estimates the actual number of COVID-19 cases in Tunisia as 2555 cases. This paper analyses the evolution of the epidemic in Tunisia using population dynamics with an SEIR model combining susceptible cases *S*(*t*), asymptomatic infected cases *A*(*t*), reported infected cases *V*(*t*), and unreported infected cases *U*(*t*). This work measures the basic reproduction number 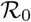, which is the average number of people infected by a COVID-19 infected person. The model predicts an 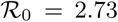. Strict containment measures have led to a significant reduction in the reproduction rate. Contact tracing and respect for isolation have an impact: at the current time, we compute that Tunisia has an 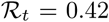 (95% CI 0.14-0.70). These values depend on physical separation and can vary over time depending on the management of suspicious cases. Their objective estimation and the study of their evolution are however necessary to understand the pandemic and to reduce their unintended damage (due to an absence of symptoms, or the confusion of certain symptoms with less contagious diseases, or unavailable or unreliable tests).

## 1 Introduction

Given the fragile health systems in most countries, we studied the data available in Tunisia as well as their possible evolution. This work presents a methodology for tracking the epidemic’s numbers. It is based on mathematical models, starting from a simple multiplication via the mortality rate [1], to a study of ordinary differential equations [13] and also to a simulation based on the Bayes theorem for the derivation of the daily reproduction rate [4]. We used a deterministic SEIR model. The main objective of this work is the estimation of the average number of infections one case can generate throughout the infectious period. It is the basic and effective reproduction number of an infectious agent.

The COVID-19 morbidity rate in Tunisia, that is, the ratio of the number of people infected to the total Tunisian population, is around 8.73 per 100 000 population. It is the number of infected cases or incidence. The death rate representing the number of people who died from COVID-19 in Tunisia is 0.4 per 100 000 people. The real indicator of the danger of the epidemic is the fatality rate, which represents the proportion of deaths compared to the total number of infected cases. The fatality rate for COVID-19 in Tunisia is 0.044 per 100 000 people. These values are temporary and incomplete. They were based on the tests carried out (the polymerase chain reaction, denoted by PCR, has a sensitivity of 70% [7]). The number of deaths is recorded in hospitals and does not include deaths at home (therefore of cases not reported). But these figures remain an indicator to contain the outbreak and prepare for targeted containment or release of confinement.

We use a set of reported data to model the epidemic in Tunisia. It is shared by the Tunisian Ministry of Health and it represents the epidemic transmission in this country. The first case was detected on March 5, 2020. Forty-five dead were reported on May 11, 2020, with 1032 total number of infected cases: the dataset is available in Table 2, and Fig. 8 gives the geographical distribution by Tunisia’s regions. The three phases of COVID-19 epidemics as defined in [11] can be decomposed as a linear phase, exponential growth, and decreasing stage. The linear growth in the number of reported cases (from March 5 to March 24) is where the number of daily reported cases is almost constant day after day. The second phase of the epidemic corresponds to an exponential increasing phase, it starts on March 25, 2020. The third phase of the epidemic starts on April 17, 2020. It corresponds to a time-dependent exponentially decreasing transmission rate, due to major public interventions and social distancing measures.

In practice, epidemiological data typically permit the estimation of the *effective reproduction number* 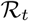. If the *basic reproduction number* 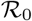 is greater than 1, countries need to take action because the progression of the virus could lead to an exponential phase. If it is less than 1 then new cases per day go to zero. Changes in transmission speed over time are recommended to bring it down the critical threshold of 1. We address the following fundamental issues concerning this epidemic in Tunisia: How will the epidemic evolve concerning the number of reported cases and unreported cases? How will the number of unreported cases influence the severity of the epidemic? What is the effect of the massive containment policies (decided on March 25, 2020, in Tunisia) on the epidemic? To answer these questions, we developed mathematical models that recover from data of reported cases and the number of unreported cases for the COVID-19 epidemic.

## 2 Methods

The average number of people contaminated by a COVID-19 infected person is central in studying epidemics. This number is denoted by 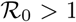: if it is less than 1, a patient infects on average less than one individual, and the disease disappears from the population over time. Conversely, if 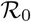, the disease can spread in the population and become epidemic. This number, therefore, makes it possible to determine if the epidemic can spread in the population, at what speed (doubling time), and with what magnitude. Its calculation requires the use of more or fewer complex models.

Compartmental models were created in the 1930s by Kermack and McKendrick [5]. The principle is to divide the population into epidemiological classes such as those susceptible to infection, those who are infectious, and those who have recovered and acquired immunity or are deceased. The state of the epidemic is determined by knowing the size of these three classes of the population. At each point in time *t*, we can consider the number *S*(*t*) of susceptible people, the number *I*(*t*) of infected people, and the number *R*(*t*) of recovered (and therefore immune) or deceased people. Each of these quantities is a function of time t, which we cannot measure at all times, but it is measured regularly at certain times (for example daily or hourly), and these measures provide both an abstract model of this function and a daily assessment of the epidemic. The functions *S*(*t*), *I*(*t*) and *R*(*t*) are linked to each other bylaws that describe how they influence each other. For example, their sum *S*(*t*) + *I*(*t*) + *R*(*t*) represents the total population which therefore remains constant over time.

The SEIR model is a little more elaborate. It is obtained from the SIR model by adding a new epidemiological class to take into account the duration of incubation, namely exposed people (infected non-infectious) who are therefore not contagious, represented by the function *E*(*t*). The purpose of the model is to predict forward in time the future number of cases in a time-line of the epidemic from early reported case data. A typical SEIR (susceptible, exposed, infectious, removed) model is given in [10]. Assume that infected individuals were not infectious during the incubation period [25]. Assume population growth rate and death rate are zero. Assume people exhibit consistent behaviours before and during epidemic phase [26]. Assume no quarantine or other mitigation intervention is implemented. All these assumptions directly influence the model, which is dependent on several parameters. See [20] for a more precise idea of the evolution of these assumptions. Sophisticated SEIR/SIRU models are developed in [21], but we choose to follow the approach of [11] to have a starting point of view of the epidemic. It produces a result closer to the realistic basic reproductive number 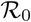.

The study we are presenting is based on a refined SEIR model, tested at the beginning of the COVID-19 pandemic in China or France [13], and in Palestine [2], which takes into account three classes of infected populations represented by functions *A*(*t*), *U*(*t*) and *V*(*t*). Any of these infected populations can transmit the virus to susceptible people *S*(*t*). The first class, represented by the function *A*(*t*), is made up of people infected but who do not know it, called asymptomatic. The other infected people, called symptomatic, are divided into two classes represented by the functions *U*(*t*) and *V* (*t*). The function *U*(*t*) represents symptomatic people not listed by the public authorities. They are infected people with symptoms but who do not know they are carrying the virus. They are therefore not officially recognised as infected (either because the COVID-19 test turned out to be negative, or because they simply escaped the various checks put in place). The population of those who are symptomatic, infected, and who tested positive for the virus are represented by the function *V* (*t*). The other laws which describe the evolution of the epidemic can be represented by the following diagram in Fig. 1:

**Fig. 1.**
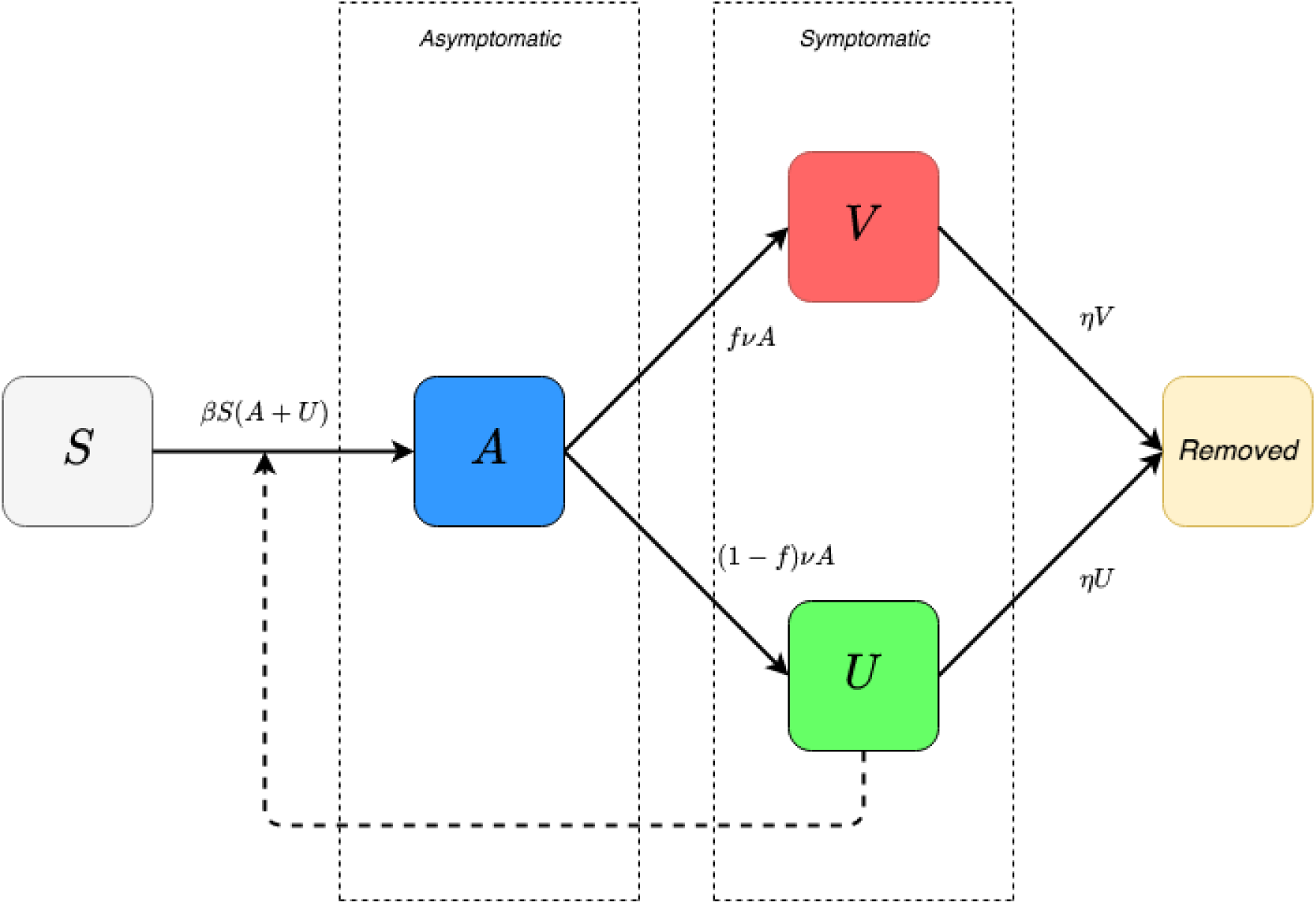
Diagram flux: refined SEIR model

The infected people are thus divided into asymptomatic *A*(*t*) and symptomatic *U*(*t*) and *V*(*t*). Only those infected who have tested positive for the virus, represented by the function *V*(*t*), are known to the public authorities. These are the ones that appear in the daily reports of the epidemic. However, the three population classes *A*(*t*), *U*(*t*) and *V*(*t*) are carriers of a highly contagious infection. Draconian control is applied to class *V*(*t*) to avoid contact with class *S*(*t*) of healthy people who are likely to be infected. However, the latter can be infected by both classes *A*(*t*) and *U*(*t*), with a transmission rate *β*: it represents the rate of susceptible people who become infected when in contact with contaminated persons. In this refined model, there are therefore *βS*(*t*)(*A*(*t*) + *U*(*t*)) newly infected people. The model also takes into account the number of days *n* from which symptoms appear on carriers of the virus (estimated at 7 days for COVID-19). After this period of *n* days, only the proportion *f* of asymptomatic people become symptomatic and test positive. They are added to the class *V*(*t*). The rest of the asymptomatic population, namely (1 − *f*)*A*(*t*), escapes the mesh of control and is added to the class *U*(*t*). Every day, class *A*(*t*) therefore contributes to class *V*(*t*) by *fA*(*t*)/*n* people and to class *U*(*t*) by (1 − *f*)*A*(*t*)/*n* people. Finally, part of the symptomatic *U*(*t*) and *V*(*t*) is removed after a certain number of days with a rate *η*. We obtain, by noting the inverse of *n* by *ν*:

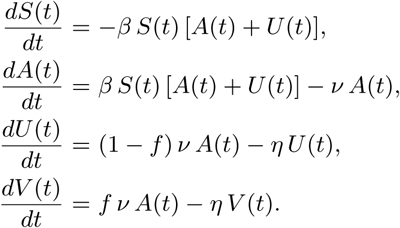

This system is supplemented by initial data *S*(*t*_0_) = *S*0 > 0, *A*(*t*_0_) = *A*_0_ > 0, *V*(*t*_0_) = 0 and *U*(*t*_0_) = *U*_0_ ≥ 0, where, *t*_0_ is the beginning date of the epidemic, *t* ≥ *t*_0_ is time in days. The parameters are listed in Table 1.

**Table 1.** Parameters and initial conditions of the model

| Symbol | Interpretation | Method |
| --- | --- | --- |
| $t_0$ | Time at which the epidemic started | fitted |
| $S_0$ | Number of susceptible at time $t_0$ (Tunisian population) | **fixed** |
| $A_0$ | Number of asymptomatic infectious at time $t_0$ | fitted |
| $U_0$ | Number of unreported symptomatic infectious at time $t_0$ | fitted |
| $\beta$ | Transmission rate | fitted |
| $1/\nu$ | Average time during which asymptomatic infectious are asymptomatic | **fixed** |
| $f$ | Fraction of asymptomatic infectious that become reported symptomatic infectious | **fixed** |
| $f\nu$ | Rate at which asymptomatic infectious become reported symptomatic | fitted |
| $(1 - f)\nu$ | Rate at which asymptomatic infectious become unreported symptomatic | fitted |
| $1/\eta$ | Average time symptomatic infectious have symptoms | **fixed** |

We assume that the removal rate *ν* is the sum of the removal rate of reported symptomatic infectious individuals, and of the removal rate of unreported symptomatic infectious individuals due to all other causes, such as mild symptoms, or other reasons. The cumulative number of reported symptomatic infectious cases at time *t* is denoted by CR(t). We assume that CR(t) has the following special form:

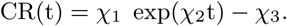

We obtain the model starting time of the epidemic *t*_0_:

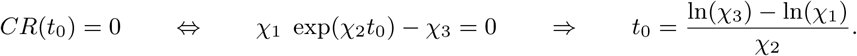

We fix *S*_0_ to 11 818 619, which corresponds to the total population of Tunisia. We assume that the variation in *S*(*t*) is small during the period considered, and we fix *ν*, *η*, *f*. We can estimate the parameter *β* and the initial conditions *U*_0_ and *A*_0_ from the cumulative reported cases CR(t). We then construct numerical simulations and compare them with data. We obtain *A*(*t*) = *A*_0_ exp(χ_2_(*t* − *t*_0_)) and *A*_0_ = χ_2_/(*fν*). We must have *U*(*t*) = *U*_0_ exp(χ_2_(*t* − *t*_0_)). So, by substituting these expressions into previous identities, we obtain: χ_2_ *A*_0_ = *βS*_0_ (*A*_0_ + *U*_0_) − *νA*_0_, χ_2_ *U*_0_ = (1 − *f*)*ν I*_0_ − *ηU*,

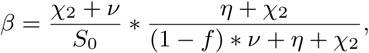

and *U*_0_ = *A*_0_((1 − *f*)*ν*)/(*η* + χ_2_). We fix *β* such that the value χ_2_ becomes the dominant eigenvalue of

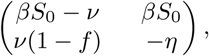

and (*A*_0_,*U*_0_) is the positive eigenvector associated with this dominant eigenvalue χ_2_. Thus, we apply implicitly the Perron-Frobenius theorem. Moreover, the exponentially growing solution (*A*(*t*), *U*(*t*)) that we consider (which is starting very close to (0, 0)) follows the direction of the positive eigenvector associated with the dominant eigenvalue χ_2_.

The need to impose lock-down comes from the classes of infected but unreported, that is, *A*(*t*) and *U*(*t*):

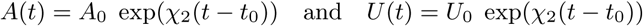

These two functions are not known at any given time *t*. Their objective estimation and the study of their evolution are however necessary to understand the pandemic and to reduce their unintended damage (due to an absence of symptoms, or the confusion of certain symptoms with less contagious diseases, or unavailable or unreliable tests).

The basic reproductive number becomes:

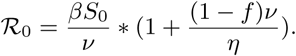

It is a single measure that does not adapt to changes in behaviour and restrictions. By definition, 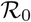 is an intrinsic property of an outbreak and does not change once calculated, as it is calculated assuming a fully susceptible population. On the other hand, the effective reproduction rate 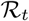 is a relevant indicator to qualify contagiousness. Every day *t*, a new case count of infected people gives us a clue about the current value of 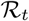. It figures that the value of 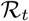 a day *t* is related to the value of 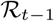 and every previous value of 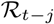 (from day *t* − *j* to *t* − 1). As a pandemic evolves, increasing restrictions change 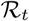. If the effective reproduction number 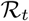 is smaller than 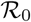, then it reduces the number of infected people, and it reduces the number of future infected people. Stopping containment, resuming international flights without any physical distancing measure, will affect 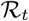 (which is likely to rise above 1).

In order to compute 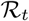 we apply a modified version of a solution used in [4] and trained by Kevin Systrom, Adam Lerer and Frank Dellaert^1^. It introduces a process model with Gaussian noise to estimate a time-varying 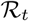. It uses Bayes’ rule to update 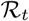 based on how many new cases have been reported each day:

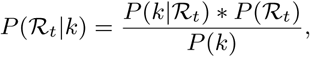

where 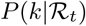 is the likelihood of seeing *k* new cases given 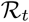 times, 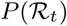 is the prior beliefs of the value of 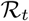 without the data, and *P*(*k*) the probability of seeing this many cases in general. To make it iterative: every day that passes, we use yesterday’s prior 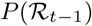 to estimate today’s prior 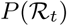. We assume the distribution of 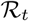 to be a Gaussian centred around 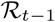, so 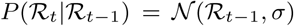, where *σ* is a hyper-parameter. So on day one:

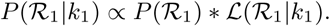

By definition, the likelihood function 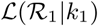 is conditioned on the observed *k*_1_ and it is a function of the unknown parameter 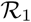. On day two:

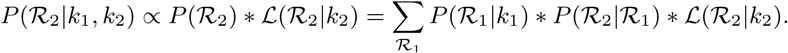

On day *t*:

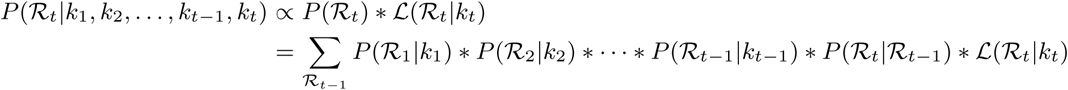

A likelihood function says how likely we are to see *k* new cases, given a value of 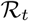: Given an average arrival rate of λ new cases per day, the probability of seeing *k* new cases is distributed according to the Poisson distribution: *P*(*k*|λ) = λ*^k^* exp(−λ)/*k*!. There’s a connection between 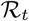 and λ:

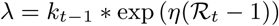

where *k_t_* is the number of infected cases at day *t*, and 1/*η* is the serial interval defined as an average residence time when infectious individuals recover or die [22].

## 3 Data description

The data available in [6] gives the number of COVID-19 positive cases per day, the number of deaths due to COVID-19, and the number of reported recoveries (see Table 2). The daily growth rate of the infected cases in Fig. 2 is given by: (*C_j_* − *C_j_*_−_*_i_)/C_j_*_−_*_i_* in day *j*, where *C_j_* is the number of reported COVID-19 cases at time *j*. The median of the daily growth rate of reported cases in Tunisia from March 5 to 11*^th^* May 2020 is equal to 0.24%. The total number of deaths is 45. It represents a fatality rate of 4.36% of total reported cases.

**Fig. 2.**
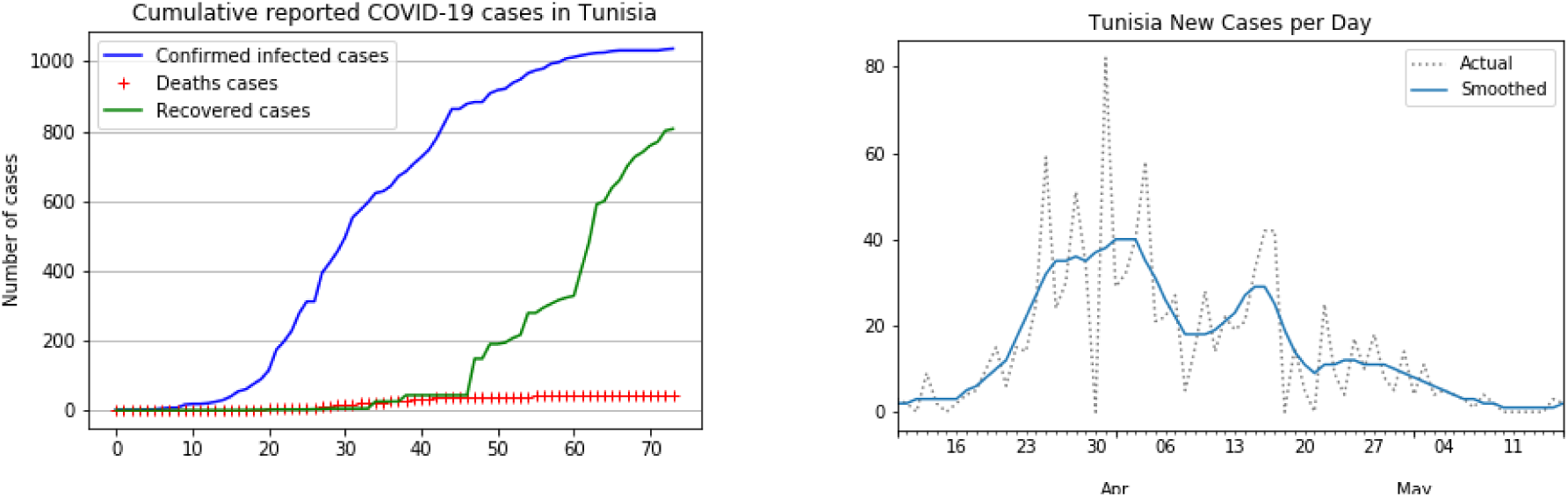
Data on reported infected cases and death cases in Tunisia were collected from January 23, 2020. The first infected case was reported on March 5. On May 11 the daily number of reported cases was zero. Based on the Ministry of Health data, we can say that the first wave of infection has passed.

Patients who die on a given day *j* were infected much earlier, so the mortality rate denominator should be the total number of patients infected at the same time as those who died [24]. To measure a death ratio, a delay of 18 days is used between the mortality number and the infected number. This delay is justified by the days between infection and death, which is about 17 to 19 days [8]. The relationships between deaths a given day *j* and the number of cases 18 days before (*j* − 18) are linear for Tunisia, as shown in Fig. 3. The mortality rate is defined by *T*_18_ = *M_j_/C_j_*_−18_ where *M_j_* is the total number of deaths at time *j*.

**Fig. 3.**
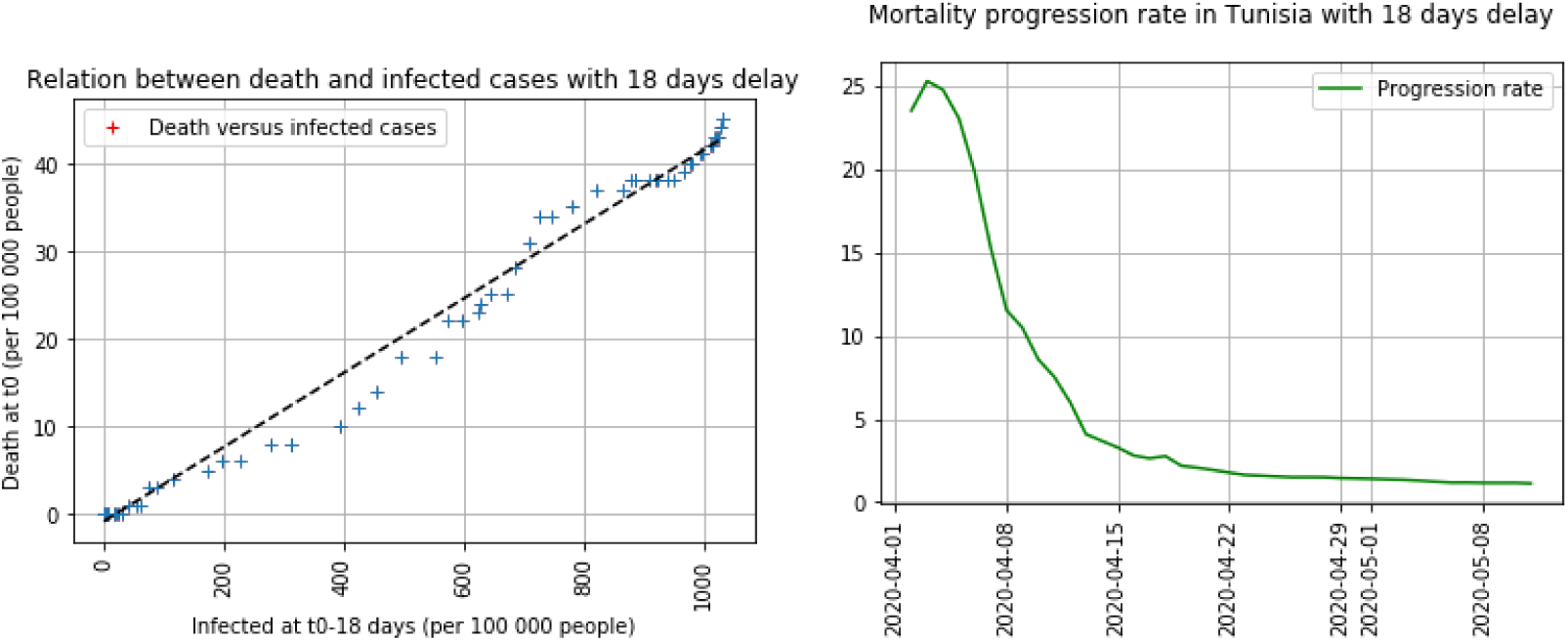
Relationships between deaths a given day j and the number of cases 18 days before *j* − 18 is linear for Tunisia. The progression rate *P*_18_ is equal to 25.28 on April 2 and decreases to 1.14 on May 11, 2020.

The median mortality rate during this period is equal to 0.12. The progression of the number of cases reported over the last eighteen days *P*_18_ is known in each country. It depends on the rate of containment and its effectiveness: *P*_18_ = *C_j_*/*C_j_*_−18_. Fig. 3 shows the progression rate of COVID-19 in Tunisia. The median of *P*_18_ is equal to 1.86 and it is used to compute the estimated number of reported and unreported cases [1]. We estimate the number of cases in Tunisia as of May 11*^th^*, 2020 by modulating the estimated mortality rate: For example, if the mortality rate is 2%, then the actual cases on May 11 is estimated at 2 555.

On the other hand, we scale the number of deaths per 100 000 population due to COVID-19 related to the number of registered infected people per 100 000 population. Tunisia appears to be doing rather well, compared to other countries [2], with a ratio of deaths per 100 000 population to infected cases per 100 000 population equal to 0.05. The incubation period for COVID-19 is very important: it is a time during which individuals have been infected but are not yet infectious themselves.

## 4 Results

The first infected case in Tunisia was documented on March 5, 2020. According to data, the time *t* = 0 will correspond to 5 March. It is used as the first day of the forecast. Fitting Tunisian data from March 25 to April 1st enables to compute the cumulative number CR(t) of reported symptomatic infectious cases at time *t*: CR(t) = χ_1_ exp(χ_2_ t) − χ_3_. We find χ_1_ = 1.885, χ_2_ = 0.209, and χ_3_ = 2.054. The straight line in the right side of Fig. 4 corresponds to *t* → ln(χ_1_) + χ_2_*t*. We first estimate the value of χ_3_ and then use a least square method (MSE) to evaluate χ_1_ and χ_2_. We observe that the data for Tunisia provides a good fit for CR(t) (explained variance score equal to 0.99).

**Fig. 4.**
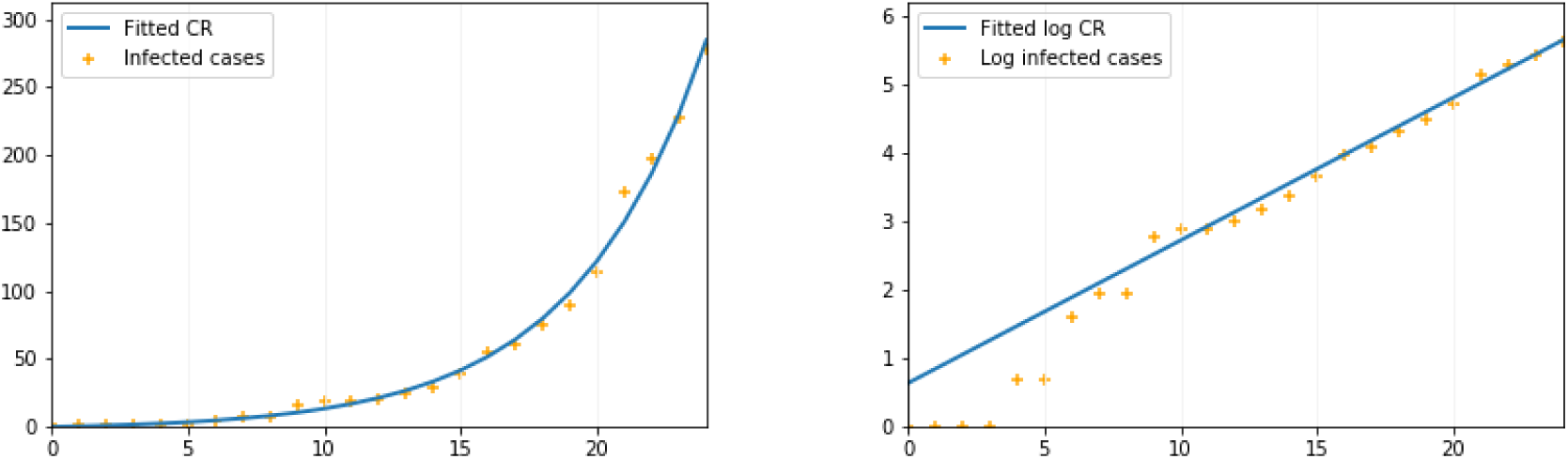
In the left side figures, the dots correspond to *t* → CR(t), and in the right side figures, the dots correspond to *t* → ln(CR(t)) − χ_3_, where CR(t) is taken from the cumulated confirmed cases.

From now on, we fix the fraction *f* = 0.6 of asymptomatic that become reported symptomatic infectious. The average time during which asymptomatic infectious cases are symptomatic is equal to 1/*ν* = 1 /7. The average time symptomatic infectious have symptoms is equal to *η* = 1/7. The values 1/*η* = 7 days and 1/*ν* = 7 days are taken from information concerning earlier coronaviruses, and are used now by medical authorities [18]. Thus, the value of *t*_0_ = 0.409 means that the starting time of the epidemic is 5 March. We obtain 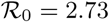, *β* = 2.76 10^−8^, *I*_0_ = 3.8, and *U*_0_ = 0.3. We ran numerical simulations and compare them with data. In the following, let take into account the fact that very strong isolation measures have been imposed for all Tunisia since 25 March. Specifically, since March 12, families in Tunisia were required to stay at home. To take into account such a public intervention, we assume that the transmission of COVID-19 from infectious to susceptible individuals stopped after 25 March. Therefore, we consider the following model: for *t* ≥ *t*_0_, *β*(*t*) = *β* if *t* ∈ [*t*_0_, 25] and *β*(*t*) = 0 for *t* > 25.

For a total population *N* equal to 11 818 619, we suppose that *S*_0_ = 11.8110^6^ is the susceptible population to be infected by COVID-19. Without these strict control policies, the number of infected cases certainly would have been different. Fig. 5 shows the number of *t* → CR(t), the unreported cases *t* → *U*(*t*), and the data corresponding to the confirmed cumulated cases for Tunisia, without containment measures.

**Fig. 5.**
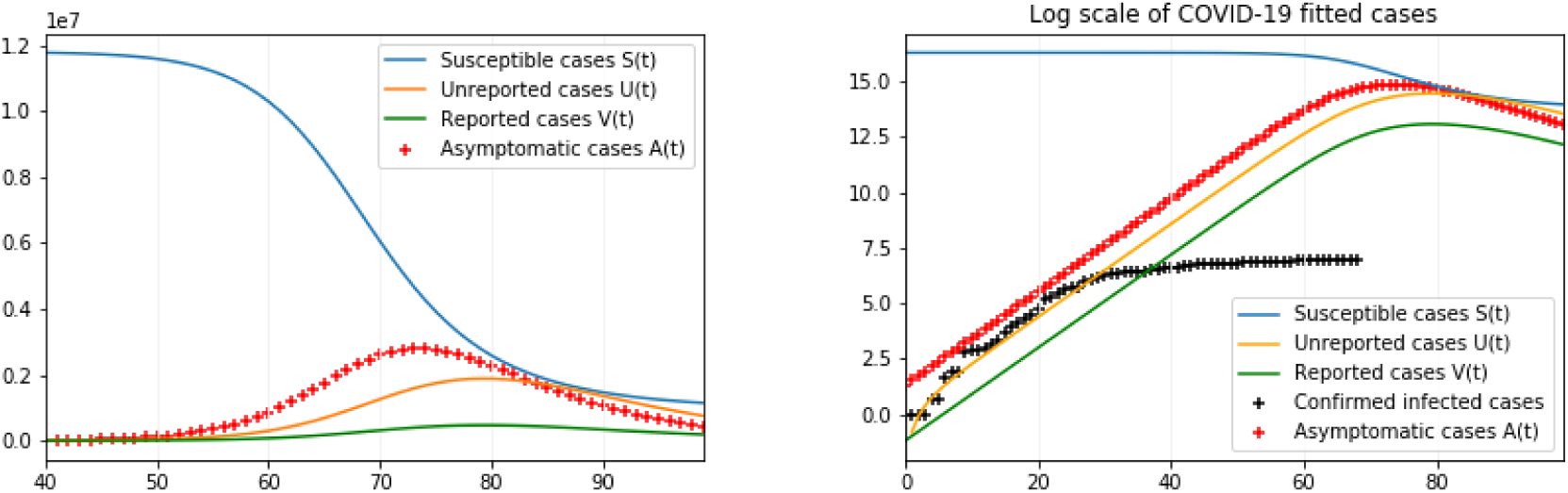
We use χ_1_ = 1.885, χ_2_ = 0.209, χ_3_ = 2.054, *t*_0_ = 0.409 and *S*_0_ = 11.8110^6^. Which gives 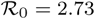, *β* = 2.76 10^−8^, *I*_0_ = 3.8, and *U*_0_ = 0.3. In the right figure, the number of symptomatic reported cases if containment has not occurred can be estimated to 472 457 i.e. 4% of the total population of Tunisisa. They would have been infected with more or less severe symptoms.

The reproduction number 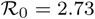 is quite high and the damage of COVID-19 came from the virulence of the symptoms. In the literature, the value of 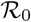 varies from 1.4 to 3.9 [17]. A more sophisticated model is developed in [3] to examine the effects of an outbreak with control policies, it gives an 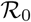 in the same range. The measure of the effective reproduction number 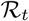 under the current severe containment conditions in Tunisia, using the Bayes’ theorem, gives 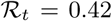 with CI 0.14 - 0.70 at May 11, 2020 (for an optimal *σ* = 0.2 and *η* = 1/7). Fig. 6 shows that it leads to small outbreaks that eventually become extinguished. To avoid overfitting on any one state, we choose the σ that maximizes *P*(*k*) over every state. To do this, we add up all the log-likelihoods per state for each value of *σ* then choose the maximum: Fig. 7 indicates the optimal result.

**Fig. 6.**
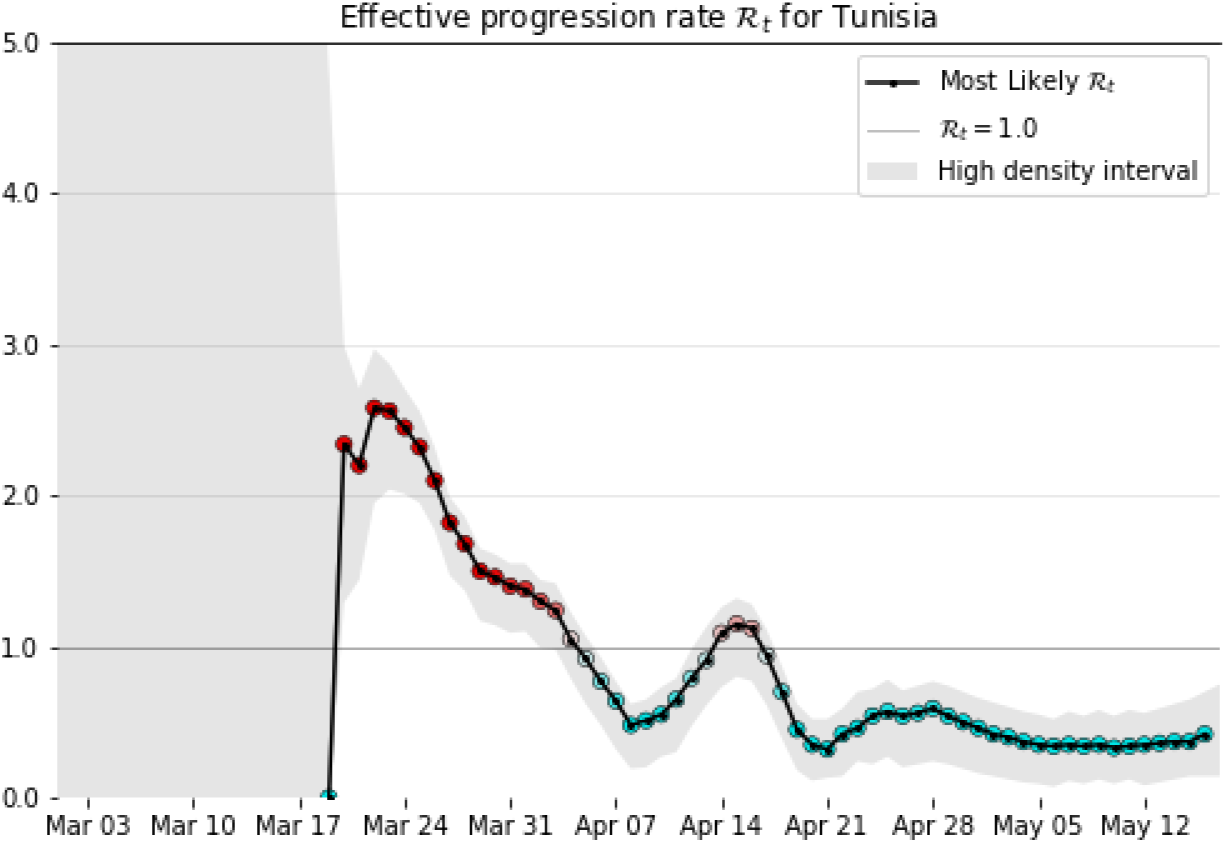
The variance *σ* is optimised on smoothed daily data. When *σ* = 0.20 and *η* = 1/7, 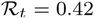 (mean CI 0.14-0.70). These results are quite unstable and are highly dependent on the number of cases detected. The downward trend could change at any time, especially as populations change their behaviour.

## 5 Discussion

The current emerging trend in mathematical epidemiology towards real-time predictive methods will allow for a shift to more quantitative surveillance and prevention policy at the earliest stages of the epidemic.

This will result in more consistent and expanded surveillance of emerging infectious diseases and improved design of health interventions and logistical allocations as epidemics develop. An epidemic outbreak of a new human coronavirus COVID-19 occurred in Tunisia. The unreported cases and the disease transmission rate are useful information. We estimate an actual number of infected cases in Tunisia based on the 18-day effect from infection to death and a mortality rate equal to 2%. We find that the number of cases on May 11 is at least equal to 2 555 infected cases, the worst case is about 4% of the population. In fact, according to the SEIR model with asymptomatic and symptomatic compartments, where the transmission rate *β* = 2.76 10^−8^, the total number of reported cases is 472 457 population near the turning point May 15. On May 11, the number of cases was 1032 infected, 45 dead, and a total population of 11 818 619. The effective production rate puts Tunisia in the leading of its neighbours. It shows very good management of the epidemic by Tunisia, especially on the number of deaths, Recovery, and Infected cases.

The estimation of 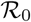 is achieved by fitting to the data (during the exponential period). This value may differ due to acquired immunity and other factors. The reproductive number 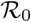 is equal to 2.73 for a fixed fraction *f* = 0.8 of symptomatic cases that are reported and for a removal rate *ν* = 7. Effective reproduction number 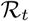 is given by matching the data, day by day, using Bayes’ theorem. On May 17, 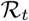 is equal to 0.42 (95% CI 0.14-0.70) where *η* =1/7 and *σ* = 0.2. The effective reproduction number 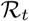 has reduced to less than 1: public measures were taken, such as isolation, quarantine, and public closings, strict travel restrictions, face mask for everyone, and reliable targeted testing. These measures exacerbate the spread of this disease and mitigate the final size of the epidemic [21]. It is contained in Tunisia, provided that quality of care and social distancing measures are in place. This protection may involve several factors, such as genetic predispositions, the immune status of populations, the history of vaccinations, and in particular what is known as *training immunity*, which is currently being discussed when BCG vaccination is involved [15]. Compared to other countries, there is a certain level of protection that must be understood (in the biological and immunological sense) to take advantage of it for better control of this disease or others that will arise. Finding a vaccine, testing the miracle cure would reduce evolution.

In Tunisia, there is strict containment, but also contact tracing and respect for isolation: 24% of the cases are foreign and the rest are domestic^2^. The reopening of the borders without severe distancing measures could have an impact and consequences for the epidemic outcome: Tunisia is heavily exposed from different Chinese airports (Guangdong province and weakly to Zhejiang province) as the main source of entry risk [9]. Several infection waves are expected, and they could be spread over several years [16]. The mathematical models outlined in this research could help anticipate the damage to the health care system that they could cause promptly. Using models applied to hospital and death data, further work will be carried out to estimate the impact of containment, as well as the current immunity of the population [19]. In terms of public health implications, to stop the outbreak, at least 58-76% of transmissions should be blocked. And we will no longer talk about this pandemic when 2/3 of the population will have been in contact with the virus. In the meantime, confinement could become a toxic situation for our democracies: For societies like Tunisia the ease with which entire sections of the population living in precarious conditions are endangered [23]. The extraordinary real-time media coverage of the progression of this pandemic has weighed heavily on the behaviour of all concerned, not only ordinary people but also politicians and even scientists [14]. The mathematics of emergency medicine can yield very relevant results.

## Data Availability

All data used is available.

https://github.com/CSSEGISandData/COVID-19

Ines Abdeljaoued-Tej, mathematician, Higher School of Statistics and Information Analysis, University of Carthage, and BIMS Laboratory, LR16IPT09, Institut Pasteur de Tunis, University of Tunis El Manar, Tunisia.

## Appendix

**Fig. 7.**
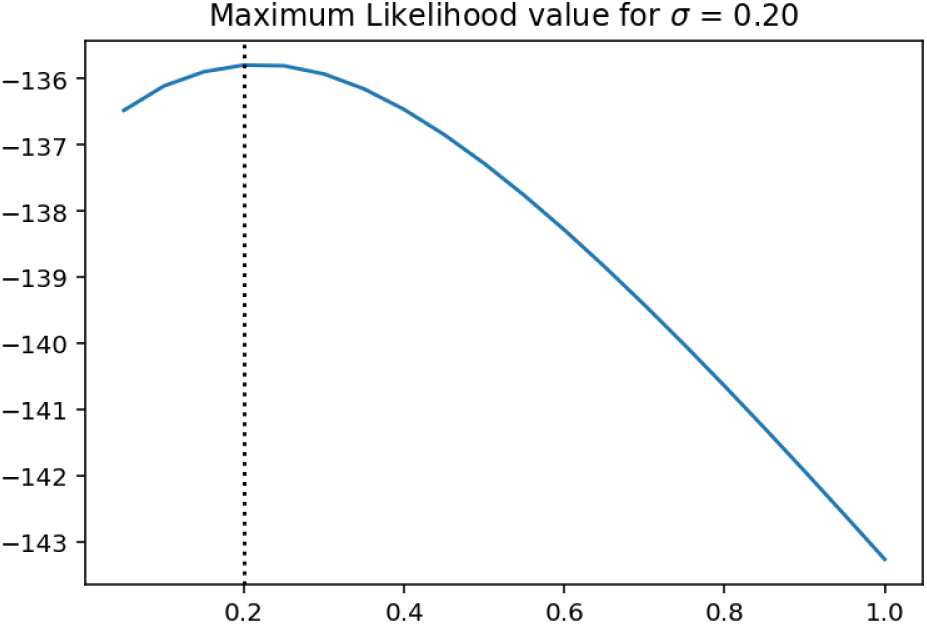
The variance *σ* is optimised on smoothed daily data from March 18 to May 17, 2020. When *η* = 1/7, we obtain the optimal value *σ* = 0.2. This result is highly dependent on the input smoothed data.

**Table 2:** The total number of infected, deaths, and recovery cases from March 3 to May 11, 2020, in Tunisia.

| Begin of Table 2 |  |  |  |
| --- | --- | --- | --- |
| Date | Infected | Deaths | Recovered |
| 3/4/20 | 1 | 0 | 0 |
| 3/5/20 | 1 | 0 | 0 |
| 3/6/20 | 1 | 0 | 0 |
| 3/7/20 | 1 | 0 | 0 |
| 3/8/20 | 2 | 0 | 0 |
| 3/9/20 | 2 | 0 | 0 |
| 3/10/20 | 5 | 0 | 0 |
| 3/11/20 | 7 | 0 | 0 |
| 3/12/20 | 7 | 0 | 0 |
| 3/13/20 | 16 | 0 | 0 |
| 3/14/20 | 18 | 0 | 0 |
| 3/15/20 | 18 | 0 | 0 |
| 3/16/20 | 20 | 0 | 0 |
| 3/17/20 | 24 | 0 | 0 |

| Continuation of Table 2 |  |  |  |
| --- | --- | --- | --- |
| Date | Infected | Deaths | Recovered |
| 3/18/20 | 29 | 0 | 0 |
| 3/19/20 | 39 | 1 | 0 |
| 3/20/20 | 54 | 1 | 0 |
| 3/21/20 | 60 | 1 | 0 |
| 3/22/20 | 75 | 3 | 1 |
| 3/23/20 | 89 | 3 | 1 |
| 3/24/20 | 114 | 4 | 1 |
| 3/25/20 | 173 | 5 | 2 |
| 3/26/20 | 197 | 6 | 2 |
| 3/27/20 | 227 | 6 | 2 |
| 3/28/20 | 278 | 8 | 2 |
| 3/29/20 | 312 | 8 | 2 |
| 3/30/20 | 312 | 8 | 3 |
| 3/31/20 | 394 | 10 | 3 |
| 4/1/20 | 423 | 12 | 5 |
| 4/2/20 | 455 | 14 | 5 |
| 4/3/20 | 495 | 18 | 5 |
| 4/4/20 | 553 | 18 | 5 |
| 4/5/20 | 574 | 22 | 5 |
| 4/6/20 | 596 | 22 | 5 |
| 4/7/20 | 623 | 23 | 25 |
| 4/8/20 | 628 | 24 | 25 |
| 4/9/20 | 643 | 25 | 25 |
| 4/10/20 | 671 | 25 | 25 |
| 4/11/20 | 685 | 28 | 43 |
| 4/12/20 | 707 | 31 | 43 |
| 4/13/20 | 726 | 34 | 43 |
| 4/14/20 | 747 | 34 | 43 |
| 4/15/20 | 780 | 35 | 43 |
| 4/16/20 | 822 | 37 | 43 |
| 4/17/20 | 864 | 37 | 43 |
| 4/18/20 | 864 | 37 | 43 |
| 4/19/20 | 879 | 38 | 43 |
| 4/20/20 | 884 | 38 | 148 |
| 4/21/20 | 884 | 38 | 148 |
| 4/22/20 | 909 | 38 | 190 |
| 4/23/20 | 918 | 38 | 190 |
| 4/24/20 | 922 | 38 | 194 |
| 4/25/20 | 939 | 38 | 207 |
| 4/26/20 | 949 | 38 | 216 |
| 4/27/20 | 967 | 39 | 279 |
| 4/28/20 | 975 | 40 | 279 |
| 4/29/20 | 980 | 40 | 294 |
| 4/30/20 | 994 | 41 | 305 |
| 5/1/20 | 998 | 41 | 316 |
| 5/2/20 | 1009 | 42 | 323 |
| 5/3/20 | 1013 | 42 | 328 |
| 5/4/20 | 1018 | 43 | 406 |
| 5/5/20 | 1022 | 43 | 482 |
| 5/6/20 | 1025 | 43 | 591 |
| Date | Infected | Deaths | Recovered |
| 5/7/20 | 1026 | 44 | 600 |
| 5/8/20 | 1030 | 45 | 638 |
| 5/9/20 | 1032 | 45 | 660 |
| 5/10/20 | 1032 | 45 | 700 |
| 5/11/20 | 1032 | 45 | 727 |
| End of Table 2 |  |  |  |

**Fig. 8.**
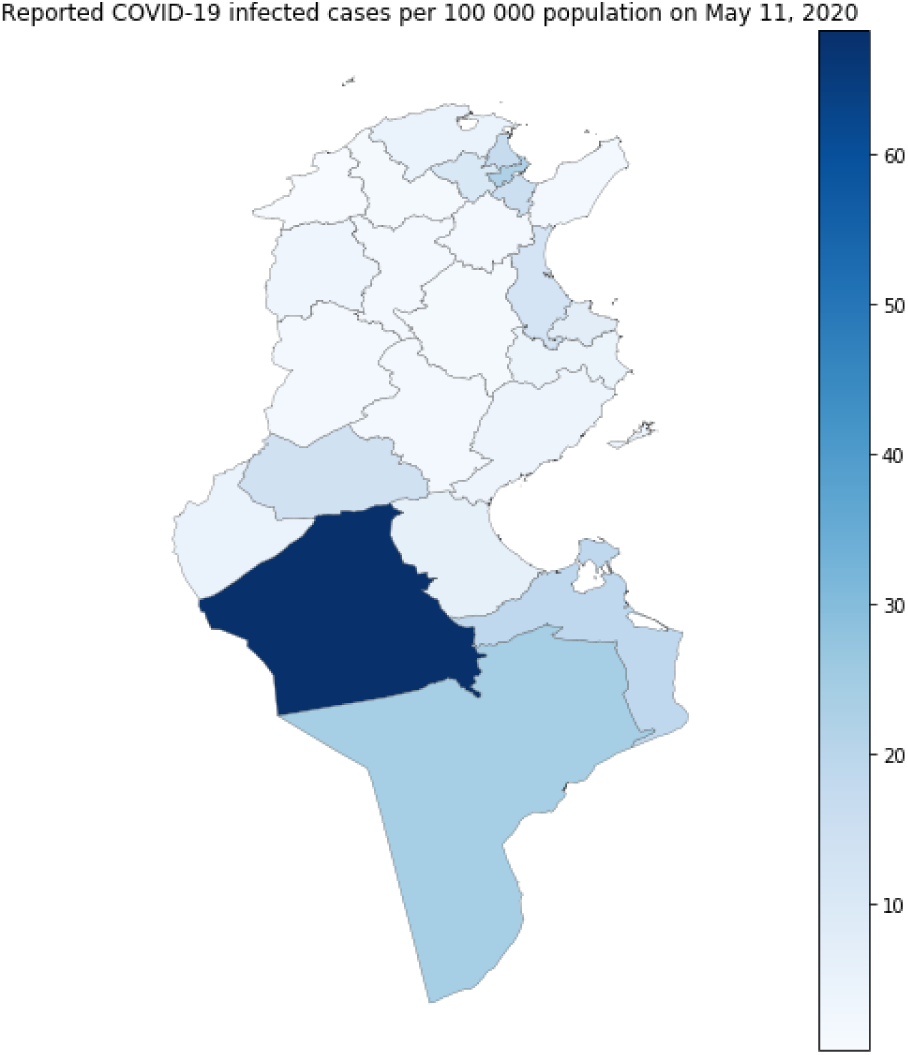
Distribution of infected cases in the 24 Tunisian regions, on May 11, 2020. The most affected regions are Kebili (south-west: 68.17 per 100 000 population), Tataouine in the south (24.09 per 100 000 population), and Tunis (22.06 per 100 000 population). Jendouba (north-west) is the least affected with 0.25 per 100 000 population.

## Footnotes

1 https://github.com/k-sys/covid-19/blob/master/Realtime%20R0.ipynb

2 https://covid-19.tn/

